# Why did you use that test? Exploring speech-language pathologists clinical decision-making in bilingual language and literacy assessment

**DOI:** 10.1101/2024.12.17.24319113

**Authors:** Emily Wood, Mariya Kika, Olivia Daub, Monika Molnar

## Abstract

**Purpose:** Our overarching goal is to advance our understanding of clinical decision-making processes in bilingual language and literacy assessment. When evaluating bilingual children, speech-language pathologists (SLPs) use static norm-referenced assessments (SAs) developed for English monolinguals more frequently than less biased dynamic assessments (DAs). To date, no research has considered why SLPs use SAs over DAs or examined SLPs’ conceptualization of validity beyond knowledge of psychometrics. In this study we explore factors that affect SLPs’ choice and use of assessments and how clinicians conceptualize and employ validity through the lens of modern validity frameworks.

**Method:** Canadian SLPs (n=21) participated in semi-structured interviews, using a guide informed by the Theoretical Domains frameworks and Kane’s Validity framework. Reflexive thematic analysis was used to generate themes.

**Results:** Clinicians rarely report using “dynamic assessment” but did “assess dynamically” by incorporating teaching in testing. When assessing oral language, SLPs acknowledged that using SAs with bilinguals may be inappropriate, but that they continue to do primarily because scores from these measures are necessary for diagnosis and accessing services. To contend with this friction between clinical beliefs and workplace requirements, most SLPs report caveats alongside SA scores SAs to contextualize findings. Though individual clinical knowledge of psychometrics and validity in assessment varies, systemic issues play a key role in perpetuating current assessment practices with bilinguals. Finally, bilingual literacy assessment practices differ. Clinicians use a wider variety of assessments and rely less on scores to achieve desired outcomes for students.

**Conclusion:** Clinical decision-making in bilingual language and literacy assessment is influenced by both individual and contextual factors. Accordingly, efforts to shift practice patterns cannot solely focus on individual clinical knowledge but must also examine and address these systemic issues.

## Introduction

Language and literacy skills are essential for academic success, social participation and professional mobility (Ritchie & Bates, 2013; Heisz et al., 2016). However, difficulties with these skills are common. In 2022, Statistics Canada reported that approximately 49% of adults scored below high-school literacy levels and 1 in 6 adults could not pass a basic literacy test (Statistics Canada, 2022a). Furthermore, approximately 7.5% of grade 1 children meet the diagnostic criteria for developmental language disorder; a lifelong condition affecting the ability to use and understand language (Norbury, et al., 2016). Individuals with language and reading-related difficulties may require clinical intervention from professionals like speech-language pathologists (SLPs) to achieve their full potential. These interventions are most effective when provided early on in a child’s development (Fricke et al., 2013; O’Connor et al., 2010). The importance of valid language and literacy assessment as a first step in identifying those who require language and literacy support cannot be understated.

SLPs are uniquely qualified to evaluate language and literacy skills (American Speech-Language Hearing Association (ASHA), 2016.; Speech-Language Pathology and Audiology Canada (SAC), 2023). The importance of valid language and literacy assessment cannot be understated. They conduct assessments to determine which children require support, and the nature or level of therapy necessary. In school-aged populations, assessment outcomes are routinely used to inform diagnoses of developmental language disorder or dyslexia; to qualify children for funding or services from agencies, states and provinces; to determine whether students meet criteria for alternative class placements; to streamline students into special education services; and to determine whether a child will receive intervention (ASHA, 2004).

SLPs have many assessment tools at their disposal to evaluate these skills. These assessments vary in their purpose, what skills they evaluate, who they can be used with, and the outcomes they provide. When choosing an assessment, it is important for clinicians to consider whether the selected tool is designed to meet the *purpose* of the evaluation; whether it has adequate *psychometric properties* relevant to that purpose; whether it is suitable to be used with the client’s *population* and whether it assesses the *skills* of interest (American Educational Research Association et al., 2014; Daub et al., 2021). As an example, a clinician evaluating whether a seven-year-old monolingual English-speaking child meets diagnostic criteria for a developmental language disorder, might elect to use a norm – referenced assessment such as the Clinical Evaluations of Language Fundamentals-5 (CELF-5, Wiig et al., 2013). Using this test (in conjunction with other information sources about development, background and language skills in context) would be a valid decision in this situation because (i) the test was developed for the *correct purpose* (e.g., making diagnostic decisions about language disorders); (ii) it has adequate diagnostic accuracy (e.g., sensitivity and specificity values greater than 80% at a cutoff of 1.33 standard deviations below the mean); (iii) it was developed for and normed on the correct population (e.g., monolingual English-speaking children between ages 5-21) (CELF-5, Wiig et al., 2013); and (iv) it evaluates the *skill(s) of interest* (e.g., a variety of oral language skills). However, this does not mean this test is inherently valid. In fact, there are myriad ways this measure can be used that are invalid. For example, it would be invalid to use this test for the *purpose* of monitoring a child’s progress on learning grammatical morphemes, as it was not designed to do so and does not have adequate *psychometrics* for this purpose. It would also be invalid to use this test to evaluate a child’s early decoding skills, as this test does not comprehensively evaluate *key skills* of interest such as phonological awareness or phoneme-grapheme correspondence. Lastly, it would be invalid to use this test to evaluate a bilingual child’s oral language skills, because they are not part of the *population* used to develop the assessment. This example highlights a key tenet of modern validity frameworks such as those proposed by Messick (1993) and Kane (2006): it is the decisions clinicians make about how to use tests that can be valid or invalid, not the tests themselves. Unlike traditional frameworks, (e.g., Cronbach, 1955; Ebel, 1972) modern frameworks do not define validity as properties of a test (e.g., construct or content validity) or contend that a test can be valid or invalid. Rather, they conceive of validity as a unitary concept and define it as the degree to which the evidence supports the interpretation and use of test outcomes for a specific purpose, in a given context (Messick, 1993).

### The Critical Role of Assessment

If the decision made to use an assessment is invalid the outcome of the assessment is also likely to be invalid. Erroneous assessment outcomes can have significant negative ramifications for children. For instance, use of static, norm-referenced language assessments developed for English monolinguals that evaluate a child’s acquired linguistic knowledge, have been shown to misidentify difficulty and result in misplacement in special education programs when used with bilinguals (e.g., Samson & Lesaux, 2009; Yamasaki & Luk, 2018). This not only alters a student’s academic trajectory, but it also has implications for their social well-being and draws limited resources away from students who truly do require support (e.g., Guiberson, 2009). Unfortunately, many norm-referenced assessments of language and literacy skills available to SLPs have been developed exclusively for use with English-speaking monolingual children. This is problematic as bi/multilingual children may underperform on these tests, not because of true language or literacy challenges but because they have had different linguistic experiences and therefore have different acquired knowledge (e.g., Bedore & Peña, 2008). In short, these language and literacy assessments cannot be used to make valid decisions about bilingual children’s skills, because they are not developed and normed for a bi/multilingual population.

Fortunately, there are alternatives to these static, norm-referenced assessments. A promising approach for bilinguals is dynamic assessment (DA). In DA, the child’s ability to learn is evaluated through incorporating teaching, feedback, prompting and re-evaluation (Grigorenko & Sternberg, 1998). Research has found that DA can act as a less-biased alternative for evaluating the language and literacy skills of school-aged bilingual children for a variety of purposes; including diagnosis (e.g., Orellana et al., 2019), predicting later ability (Hunt et al., 2022), and capturing learning modifiability (Petersen et al., 2016). This is because DA minimizes the impact of prior linguistic or literacy experience on test outcomes providing a more equitable assessment (e.g., De Lamo White & Jin, 2011; Wood et al., 2024). For this reason, use of DA in bilingual assessment is recommended by regulatory bodies (e.g., ASHA, n.d., SAC, 2024).

### Current Language and Literacy Assessment Practices

Several studies have examined current language assessment practices with bilingual populations to determine what types of assessments clinicians use and how often they use them. A survey of over 400 American school-based SLPs in Michigan found that 98% of respondents used measures developed for English speaking children to evaluate the language skills of bilinguals (Caesar & Kohler, 2007). The most frequently used tests to evaluate bilingual children were the same as those used to evaluate monolinguals: omnibus language tests like the CELF (Wiig et al., 2013) and vocabulary measures like the EVT (Williams, 2018) and PPVT (Dunn, 2018). Language sampling, parent and teacher interviews, and classroom observations were used infrequently, and DA was not mentioned by any clinicians who participated in the survey (Caesar & Kohler, 2007). These assessment practice patterns were not affected by amount of clinical experience or the proportion of bilingual students on the clinical caseload. A follow up survey about bilingual language assessment practices found that over the subsequent ten years, SLPs had increased their use of more linguistically and culturally appropriate assessments such as language sampling, natural observations and parent interviewing (Arias & Friberg, 2017). However, less than a third reported using DA, citing barriers like lack of time, training and unfamiliarity with the approach (Arias & Friberg, 2017). Similar trends were observed in a survey of Australian SLPs (Denman et al., 2021) Canadian SLPs, though rates of use of DA were higher, with 45.9% of clinicians reporting use of the approach with bilinguals (D’Souza et al., 2012). Notably, a recent survey of American SLPs called into question how DA was being used and prioritized in the context of a comprehensive assessment. When considering best practices for multilingual populations, only 55% of clinicians rated use of DA as a high priority, and less than half reported using DA in multilingual language assessment (Nelson & Wilson, 2021). Taken together, these findings suggest that at least in some instances, clinicians are using tests that were not developed for the *population* of interest when evaluating bilinguals.

However, most of these survey studies have focused on what assessments clinicians use and how often they use them, while neglecting to address *why* clinicians choose the assessments they do in practice. At present, we understand what tests SLP use, but without asking clinicians why they use the tests they do, we cannot understand what factors lead to SLPs favouring certain assessments over others. Developing our understanding of the factors that shape clinical decision-making is critical for shifting practice patterns. For instance, without understanding *why* clinicians don’t use DA regularly, we can only guess what strategies might be useful in promoting its uptake. For example, providing training and engaging in efforts to grow awareness of DA will only shift practice if we find that clinicians do not use DA because they are unaware of it. This strategy would be ineffective if we find that clinicians are aware of it, but don’t use DA for other reasons.

### Clinical Conceptualizations of Validity Inform Understanding of Practice

Previous survey studies have examined whether validity might be one such factor that influences assessment choice and use. These surveys have primarily focused on the knowledge of psychometric properties as a proxy for clinical knowledge of validity (e.g., test reliability and validity, standard error of measurement, sensitivity and specificity etc. [e.g., Betz et al., 2013; Kerr et al., 2003]). Results of these studies indicate that SLPs feel only somewhat confident in their knowledge of test psychometrics (Kerr et al., 2003). Many SLPs were not able to define why using a test in a certain way could be considered invalid (e.g., using a test developed for diagnosis to set intervention targets). On occasion, when SLPs recognized concerns with test use or psychometrics, they reported continuing to use the tests in that way regardless (McCauley & Swisher, 1984). This suggests that other factors are at play. Examining knowledge of psychometric properties in isolation does not capture the ways different levels of evidence manifest as validity in decision-making. This is because assessments cannot be inherently valid or invalid based on these properties alone. Valid assessment means choosing the right test, for the right purpose, for the right person, and considering test outcomes and consequences.

To our knowledge, no previous studies have considered clinical decision-making through the lens of modern validity frameworks in the context of bilingual language and literacy assessment. Exploring what factors affect use of different assessments (e.g., static norm-referenced or dynamic tests) and examining how clinicians conceptualize valid assessment practices will provide a more robust understanding of clinical decision-making processes and assessment practices in bilingual language and literacy assessment. Knowledge of these factors is also necessary to design evidence-informed support for improving valid assessment practices for bilingual children (Yakovchenko et al., 2023).

### The Current Study

The current study investigates factors that affect choice and use of assessments (e.g., static norm-referenced vs. dynamic) and how clinicians conceptualize and employ validity knowledge with the goal of advancing our understanding of clinical decision-making processes in bilingual language and literacy assessment. To achieve this, we employed a qualitative approach, using semi-structured interviews. Interviews are among the most effective means of understanding “why” people act the way they do, and what underlying reasons or beliefs affect their actions and choices (Rosenthal, 2016). Unlike open-ended survey questions, interviews also allow the researcher the opportunity to ask follow-up questions, to probe for further information when required, and to clarify unclear statements. This results in a deeper understanding of perspectives of participants (Patton, 2003).

We also focus on populations (e.g., bilinguals), domains of assessment (e.g., literacy) and places (e.g., Canada) that have received less interest in prior literature. To date, no qualitative research has been conducted examining assessment practices of Canadian clinicians, and to our knowledge only one quantitative survey study has examined how frequently clinicians use certain types of assessments and what barriers they encounter in providing service to linguistically diverse clients (D’Souza et al., 2012). Consequently, very little is known about Canadian SLP bilingual assessment practices. This is relevant given that Canada is a bilingual country (English / French), with substantial linguistic diversity (Statistics Canada, 2022b). Nearly 40% of the population is bilingual, and over 200 languages are spoken across the provinces and territories (Statistics Canada, 2017). Factors that affect clinical assessment practices in Canada may also be relevant to other countries with high rates of linguistic diversity where English is the societal language, and where SLPs have similar training, education and assessment responsibilities. This includes for example, the United States, the United Kingdom and Australia (United States Census Bureau, 2022). Furthermore, most studies examining practice patterns with school-aged children, bilingual or monolingual, have focused on oral language assessment, neglecting literacy (e.g., Arias & Friberg, 2017; Caesar & Kohler, 2007). Though literacy is within the scope of SLP practice, it is possible that practices differ across language and literacy assessment (ASHA 2016; SAC, 2023). It is critical to consider literacy assessment practices as outcomes from these assessments also have academic and social implications for students.

### Research Aims

Our research aims are:

(i) to explore what factors influence SLPs’ choice and use of assessments and;
(ii) to develop our understanding of how SLPs conceptualize and employ knowledge of validity; with the goal of better understanding clinical decision-making processes in bilingual language and literacy assessment.

## Method

### Ethics

Procedures as outlined in the study protocol received approval by the University of Toronto’s Health Sciences Research Ethics Board (Protocol #44406).

### Methodological paradigm

We employ a pragmatic approach in this study. Pragmatism prioritizes the feasibility, utility, and practicality of research (Onwuegbuzie & Leech, 2005). Ontologically, pragmatism is consistent with a pluralist ontology, which acknowledges both the objective physical world and the subjective social and psychological world (Morgan, 2014). This ontological pluralism aligns with our proposed research. We contend that clinical practice is subjective, in that all clinicians have unique knowledge and lived experiences which shape their reality. However, clinicians also operate in an objective physical world, which constitutes the systems and agencies that employ them, and these objective realities may constrain or otherwise impact their practice. In terms of epistemology, pragmatism seeks to generate knowledge in whatever way(s) works to answer the question or address the problem (Morgan, 2014).

### Recruitment

Clinicians were recruited through social media posts to X, emails sent to professional organizations across Canada (e.g., to Speech-Language Pathology and Audiology Canada), via posts to community message boards and forums (e.g., SLP Facebook groups) and via direct contacts to former colleagues and employers of the first author of this paper.

## Measures

### Eligibility screener

Interested participants were prompted to fill out an eligibility screener hosted on Microsoft forms. The questions in the self-administered screener determined whether the respondent met the eligibility criteria of:

(i) being a speech-language pathologist currently practicing in Canada who
(ii) conducts language and/or literacy assessments with bilingual populations.

Participants also provided their name and email for follow-up. Informed consent was obtained at the outset of the screener to collect this personal information.

### Demographic questionnaire and consent to participate

Potential participants who met the eligibility criteria were sent a link via email to complete informed consent and a demographic questionnaire. The informed consent document provided them with information about the study, potential risks and benefits, information about where data would be stored, time required to participate and compensation. Following signature of the consent form, they were automatically directed to the demographic questionnaire, which asked questions regarding their location of practice (i.e., province and city), the type of practice in which they worked (i.e., schoolboard, private clinic), how long they had been practicing, their racial, ethnic sex, gender and linguistic identity. We collected this information to ensure that we received a diverse range of clinical perspectives from individuals across the country. All questions were open-ended.

### Interview guide

I (EW) developed the semi-structured interview guide with support from OD and MM. I considered the Theoretical Domains Framework while developing questions that would allow me to understand what factors influence the use of dynamic vs. static assessments in bilingual language and literacy assessment (Cane et al., 2012). The TDF is a framework comprised of 14 domains that can be used to identify factors that influence health professionals’ behaviour in terms of uptake of evidence-based practices (Cane et al., 2012). Examples of questions that were designed to elicit responses detailing factors influencing use of assessments include “Have you used dynamic assessments, if so, what has facilitated this / if not what has prevented you from doing so?” This question allowed clinicians to identify what impacts their use of dynamic assessment. From a TDF perspective, I hypothesized that any number of domains might impact their assessment choice. For example, the domains of *knowledge* (an awareness of DA), *skills* (an ability to use dynamic and static assessments), e*nvironmental resource*s (access and cost of various assessments), *social influence* (clinical norms in assessment practice) and beliefs *about consequences* (expected outcomes of assessments) could all play a role. In asking a broad question like “What has facilitated or prevented your use of DA?” we allowed clinicians to identify on their own any TDF domains, or other factors that impact their assessment practice. This contrasts with prior survey research which often lists potential barriers and facilitators (e.g., “lack of appropriate assessment tools for bilinguals” an environmental resource issue) and asks clinicians to rate how frequently these issues arise in their practice.

I also considered Kane’s validity framework in developing the interview guide to generate questions that would help me understand how clinicians’ conceptualization of validity and how it affects decision making processes in assessment (Kane, 2006). Like Messick (1993) Kane conceives of validity as the decisions made in assessment, rather than as a characteristic of a test (Kane, 2006). Kane outlines the types of decisions a clinician must make, and a series of inferences (e.g., scoring, generalization, extrapolation and real-life implications) that clinicians must provide evidence for or against when deciding whether a given measure is valid for use (Cook, 2015). Again, rather than asking clinicians close-ended questions about whether or how often they consider “psychometric properties” or “validity” in test selection, we posed open-ended questions like “How have you typically decided these measures are (or are not) of high quality?” “How did you decide this test was appropriate for use?” “Typically, what are the outcomes of your assessment?” These types of questions were designed to let us hear firsthand from clinicians what information they use to inform their assessment selection and how they conceptualize valid or appropriate decisions in practice.

I also developed and used a variety of types of initial and follow-up questions (e.g., task-related grand tour, contrast), to ensure we received rich responses. I considered the flow of the questions, and elected to start with broader, more descriptive questions, then to move to more sensitive topics, before transitioning back to more forward-focused, ideal future scenario questions to end the interview on a positive note. The interview guide can be accessed in the supplemental material.

### Participants

Twenty-one clinicians met the eligibility criteria for participation and responded to follow up emails to schedule an interview and participate. Twenty of the SLPs identified as cisgender women, 1 identified as a cisgender man. Their ages ranged from 27-71 with an average age of 37.5 years old. Most SLPs were Caucasian (N=13). Other reported racial and ethnic identities were Southeast Asian (N=2), Arab (N=1), Inuit (N=1), Black (N=1), South American (N=1), Indian/Caribbean (N=1) and Caucasian/Chinese (N=1). Only 4 of the SLPs were English monolinguals. The other 17 were bi/multilingual and spoke a variety of languages including French (N=12), Italian (N=3), Tagalog (N=1), Arabic (N=1), Cantonese (N=1), Russian (N=1), Hebrew (N=1), Tigrinya (N=1), Spanish (N=1) and Japanese (N=1). SLPs were from central, western and northern Canada. The largest proportion of SLPs were from Ontario (n=12), followed by Alberta (N=4), British Columbia (N=2), and Québec (N=1), Nunavut (N=1) and Yukon (N=1). Efforts were made to recruit clinicians from eastern Maritime provinces to ensure diversity of perspectives and representation. However, these efforts were unsuccessful and no SLPs from eastern provinces participated. Most SLPs worked in Urban settings (N=18) compared to rural settings (N=3), and most worked in public settings like schools or hospitals (N=12) compared to private practice (N=4). Some worked in both public and private roles (N=5). Their years of experience ranged from 2 to 30+ years. Information about the participants’ sex, language status, practice setting (urban or rural), workplace (private, public or mixed), and number of years of experience can be found in Table 1.

**Table 1.** Participant demographic characteristics.

*Participant demographic characteristics*
| **ID** | **Sex and Gender** | **Language status** | **Work -place type** | **Work context** | **Years Practiced** |
| --- | --- | --- | --- | --- | --- |
| P2 | Cisgender woman | Monolingual | Private Practice | Urban | 6 |
| P3 | Cisgender woman | Bi/multilingual | School | Urban | 9 |
| P4 | Cisgender woman | Bi/multilingual | School | Rural | 3 |
| P6 | Cisgender man | Bi/multilingual | Mixed | Urban | 23 |
| P7 | Cisgender woman | Bi/multilingual | Mixed | Urban | 3 |
| P9 | Cisgender woman | Bi/multilingual | School | Urban | 5 |
| P10 | Cisgender woman | Bi/multilingual | School | Urban | 24 |
| P11 | Cisgender woman | Bi/multilingual | Private practice | Rural | 16 |
| P12 | Cisgender woman | Monolingual | School | Urban | 30+ |
| P13 | Cisgender woman | Monolingual | School | Urban | 23 |
| P14 | Cisgender woman | Bi/multilingual | Mixed | Urban | 11 |
| P15 | Cisgender woman | Monolingual | School | Urban | 10 |
| P16 | Cisgender woman | Bi/multilingual | Mixed | Urban | 15 |

| Why did you use that test? |  |  |  |  |  |
| --- | --- | --- | --- | --- | --- |
| P17 | Cisgender woman | Bi/multilingual | Private practice | Rural | 8 |
| P18 | Cisgender woman | Bi/multilingual | School | Urban | 11 |
| P19 | Cisgender woman | Bi/multilingual | School | Urban | 9 |
| P21 | Cisgender woman | Bi/multilingual | School | Urban | 2 |
| P22 | Cisgender woman | Bi/multilingual | School | Urban | 8 |
| P23 | Cisgender woman | Bi/multilingual | Private practice | Urban | 2 |
| P25 | Cisgender woman | Bi/multilingual | Mixed | Urban | 2 |
| P27 | Cisgender woman | Bi/multilingual | School | Urban | 2 |

Mixed = both public setting (i.e., school) and private setting (i.e., private practice).
Participants were assigned codes at eligibility, and so those who did not respond or who did not consent (e.g., P5) are not included in the table as they did not participate in the study.

### Reflexivity Statements of Research Team

Each member composed a statement that summarizes their connection to this research. I (EW) am a clinical speech-language pathologist and PhD Candidate whose research focuses on dynamic assessments of early literacy skills. I am a Caucasian female, whose maternal language is English but consider myself functionally but not culturally multilingual. I have experience conducting language and literacy assessments with bilingual children. MK was a second-year speech language pathology student at the University of Toronto, working as a research assistant in the Bilingual and Multilingual Development Lab at the time of this project. She is now a practicing speech-language pathologist. She is a second-generation Gujarati-Canadian Muslim female. Her first language is English, but she learned French in school, and is receptively bilingual in Gujarati and Urdu, having grown up in a multicultural and multilingual community. OD is an Assistant Professor in the School of Communication Sciences and Disorders at the University of Western Ontario. Her research program focuses on clinical decision-making based on norm-referenced test scores, validity constructs, and integrated knowledge translation. She is a Caucasian female, raised in a monolingual English-speaking home and community. Lastly, MM is a multilingual, Caucasian, cis-gender mother and newcomer to Canada. She has more privileges and access to healthcare than most other newcomers because of her Canadian academic background. She is a professor of child language at the University of Toronto and her research focuses on bilingual language and cognition across the lifespan.

## Procedures

### Semi-structured Interviews

I (EW) conducted individual semi-structured interviews with all participants over zoom between the months of March and August 2023. Each interview lasted about an hour and was audio and video recorded. I reminded all participants of the broad study purpose at the interview’s outset, reviewed the information in the consent document they had already signed, and verbally obtained informed consent to record the interviews before beginning. All participants agreed to recording.

Before beginning interviews in earnest, I conducted one practice interview with a retired SLP who I knew personally (P1). The data collected from this interview was not included in the analysis. I did this to familiarize myself with the interview guide and the flow of questions. Following this practice interview, I adjusted some of the phrasing of the questions in the guide that were not clear based on this clinician’s feedback. I also changed how I introduced myself. I elected to refer to myself as a former schoolboard SLP and current clinician researcher, rather than simply a researcher, to minimize any potential power imbalance that may have resulted from a researcher /subject dynamic.

The semi-structured interview guide was used in all interviews. Though all questions were addressed in all interviews, the order and flow of questions was not identical across participants. In some instances, further follow-up questions were required to understand a clinician’s response, and in other cases, it was logical to jump back and forth between questions to maintain conversational flow.

### Analytic approach

I analyzed the data collected in this study using reflexive thematic analysis ([RTA] Braun & Clarke, 2023). I elected to use RTA because it is easily accessible and allows the methodological flexibility required for a pragmatic approach. In RTA, the researcher is active in the analysis and production of themes. While codes and themes generated in the analytic procedure are meant to represent the participants accounts and perspectives, they also reflect my own interpretation of the data. The reflexivity element of RTA was critical to me, given that I have lived experience as a clinician conducting language and literacy assessments with bilingual children, and as a researcher developing a literacy assessment for bilingual children. In RTA, rather than attempt to remain entirely objective, the researcher can acknowledge experience and interpret the data reflexively. Because reflexive TA can be used in many ways, it is important to explicitly state how researchers engaged in analysis. Our orientation to the data is experiential, meaning we endeavor to examine and investigate the meaning of participant experiences, rather than to interrogate them, as in a critical approach. While we did use frameworks to develop our interview guide questions, I take an inductive approach to analyzing the data, meaning that I attempted to produce codes and themes that reflect the content of the interviews, rather than trying to fit data into pre-specified frameworks or codebooks as in a deductive approach. Finally, I employed semantic coding, meaning that codes and themes represent descriptive surface-level representations (e.g., what participants verbally stated in interview.)

### Analytic procedures

I followed the six phases of reflexive thematic analysis as outlined by Braun and Clarke (2023). First, I generated a transcript of the audio and video recorded interviews and read through each transcript repeatedly to familiarize myself with the participant accounts. MK checked and edited all transcripts to ensure they were consistent with audio and video recordings. All transcripts were then uploaded into NVivo for analysis (NVivo 12, 2017). While re-reading the transcripts, I open-coded all statements that felt pertinent, interesting, valuable or informative. MK also independently coded 5 transcripts using the comment and highlight feature in Microsoft word. MK and I met weekly from May-August 2023 to discuss coding and our data interpretations. We also met monthly with the other members of the research team during this time to discuss these codes. OD and MM also read through and shared general comments through group discussion on 10 transcripts each. This was not meant to act as a reliability check or used to consolidate our codes and ideas but to facilitate an open discussion of our interpretations of the data. After two rounds of open-coding, I began collating preliminary codes (e.g., outcomes tied to test scores, no scores no respect, reports and scores have value) into higher level preliminary potential themes (e.g., scores are valuable). Themes reflect a pattern of response in the data and must be linked to the research aims. I generated several candidate themes in NVivo, but transitioned to pen and paper as I conceptualized how these themes might or might not fit together in a larger thematic map. During this process I reviewed themes and checked them against the data. I also shared thematic maps with the research team in monthly meetings. As I generated the final thematic map, I named and defined the themes and began to select illustrative quotes that best represented the themes and participant perspectives. These quotations are presented in the results section. The ellipses are used when any irrelevant information is extracted. Participants’ names are not reported.

### Trustworthiness and analytic rigour considerations

The underlying theoretical assumptions of our approach are not consistent with checking for reliability of coding through tools like a codebook or team consensus. In our view, no two researchers engaged in RTA could arrive at the same exact conclusions from this data, because we acknowledge individual realities and interpretations of each researcher. Rather, we strive to produce a trustworthy analysis that meets the criteria of being credible, transferable, dependable and confirmable (Lincoln & Guba, 1985). Credibility pertains to the notion that results are true and believable (Lincoln & Guba, 1985). To meet this criterion, we tested our interview guide prior to data collection, ensured we had the necessary knowledge and research skills to develop a guide, carry out interviews and perform reflexive TA, and had regular team debrief meetings to discuss codes, preliminary themes and maps considering multiple and diverse perspectives. Dependability in qualitative research means that the methodological procedure is clear and thoroughly documented to the extent that a separate team of researchers could repeat the study (Lincoln & Guba, 1985). To meet this criterion, we created and retained drafts of study protocols, interview guides and the first author kept a reflexivity journal documenting changes as they were made. Confirmability and transferability are related to the degree of confidence that these results could be confirmed by another research team, and the extent to which results generalize or can be transferred to other contexts (Lincoln & Guba, 1985). To meet these criteria, I kept a reflexive journal documenting my thought processes as the analysis progressed, and we kept track of participant demographic data to ensure a diverse and representative sample.

## Results

Through thematic analysis I generated five themes. Themes 1-3 correspond to objective 1, and themes 4-5 correspond to objective 2. The themes are (i) Doing assessment dynamically, (ii) One test to rule them all (iii) Scores reign supreme, (iv) Caveats and contextualizing and (v) Clinical knowledge vs. system constraints. Theme two and three both have subthemes regarding language vs. literacy assessment practices, as we noted differences between how these themes were expressed across the two domains. The overall thematic map including primary themes and subthemes is visualized in Figure 1.

**Figure 1.**
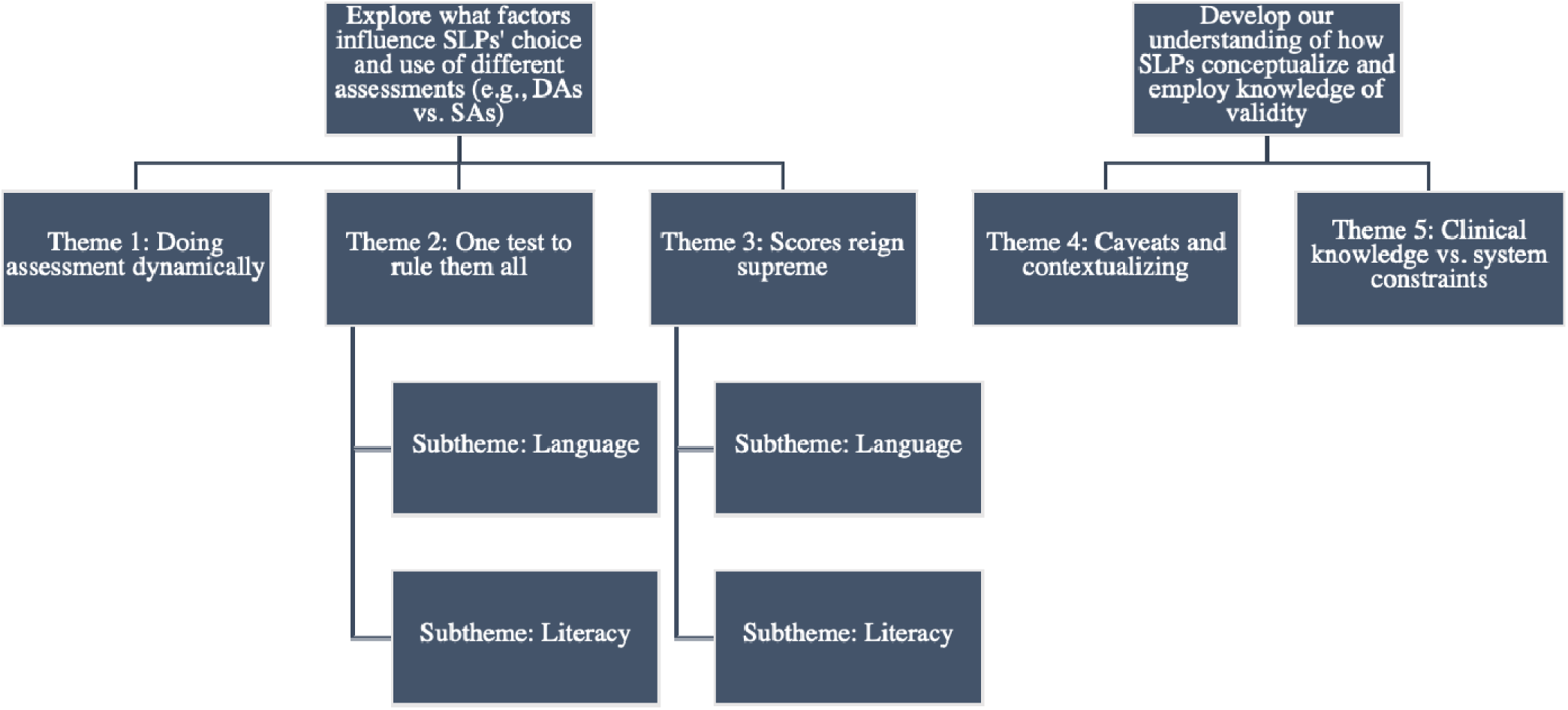
Thematic Map.

### Objective 1: Explore factors that influence clinical use of assessments (e.g., dynamic vs. static assessments) in the evaluation of bilingual children’s language and literacy skills

#### Theme 1: Doing assessment dynamically

When asked in the interviews to describe their assessment process for a hypothetical evaluation of a bilingual seven-year-old’s language and literacy skills, few clinicians confidently reported using what they referred to as “dynamic assessment” for language or literacy assessment. *“When I say dynamic assessment, I’m actually meaning like those particular assessments, like the QPS, the CUBED, the PAST, those are assessments that you can do and teach at the same time.”* (P15) However, most clinicians reported using elements or principles of a dynamic assessment approach like prompting, teaching or giving feedback in the assessment. This was true for informal language assessments like language sampling or play observation; *“I might kind of probe like a preposition, let’s say like play a game with the kiddo and kind of model like ohh let’s put the pig in the house, let’s say and maybe they put it beside and I might kind of model that with–so that’s the pretest I guess is kind of prompting for that and correct I’ll kind of model it, put the pig in the house and then at we’ll do that posttest after I teach, and then see if he can put the pig in the house after.”* (P23) for formal, standardized language assessments; *“Sometimes what I will do is you know, administer it* [CELF-5] *as it’s meant to be administered, but if the student makes an error, I give them zero for that item, but then I keep probing and just see what would they come up with if I could do that right. Like more repetitions clarification um or you know giving an example I think just sort of typical kind of scaffolding and prompting and those things that we would typically do just to see if they help.”* (P10); *“I don’t ever follow the official protocol for any of the tests. To me it doesn’t feel conducive to getting good recommendations either for support. I view testing also as a way of kind of seeing what kind of supports can be used and can be useful. So, for example like on the like vocabulary test, if students are having trouble with word finding, I’m going to adjust and I’m going to give you know clues and, and cues and see which kind of cues are going to help” (P22*) and for formal and informal assessments of early literacy skills such as a phonological awareness screener. *“If I’m doing, let’s say, phonological awareness, then I’ll kind of check like if I give like little, little support so you know, if we’re using blocks, you know, do they, does it go better?”* (P16)

Even though most clinicians identified and implemented these dynamic assessment principles into their oral language and literacy assessments, they were unsure if their approach could be considered “true” dynamic assessment. *“And like you can correct me if I’m wrong, like so dynamic assessment… I don’t know if dynamic assessment is the right word but just like giving the child the opportunity to like to learn, as opposed to just like testing.”* (P21) There appeared to be uncertainty as to whether dynamic assessment was a distinct type of assessment or a set of principles that could be applied to any assessment. *“Dynamic assessment as like a, like a principle rather than like a specific…? I’ve definitely played around with assessment materials in the test teach retest sort of style.”* (P4) Clinicians seem to have knowledge of and the ability to apply dynamic assessment principles but limited confidence in this knowledge. *“Yeah, I think my assessment is dynamic, even though it’s not like a dynamic assessment.”* (P2)

Those clinicians who reported using dynamic assessment approaches or principles, were asked follow-up questions about what facilitated their ongoing use of this practice. Clinicians reported a variety of reasons including that assessing dynamically provides valuable information for setting realistic treatment goals; *“I get more information from dynamic assessment and intervention sessions, with regards to coming up with focused and appropriate goals”* (P11) that testing dynamically gives additional information about how the child learns and responds; *“But it* [dynamic assessment] *is just to kind of see like is this kid like, do they almost have it or do they really not have it right? You know, so that kid who can get it with that little bit of prompting… Sometimes that gives me a sense of the bigger cognitive picture, right?”* (P10) and that the approach was more strengths-based as opposed to deficits-based. *“Dynamic assessment allows me to go get that information as well like, no, he can’t do this but look what he can do when I do this and stuff.”* (P16) Interestingly, few clinicians highlighted that the approach can be used to distinguish between difference and disorder in culturally and linguistically diverse populations. *“I think it just helps differentiate language difference delay or disorder or just if they’re, they just speak a different language.”* (P25)

In summary, clinicians have knowledge of, regularly implement, and see value in dynamic assessment principles but do not always define their use of these principles as “dynamic assessment,” They report that these principles are primarily useful for setting treatment objectives or recommendations rather than for differentiating disorder from language difference in bilingual populations.

#### Theme 2: One test to rule them all

##### Subtheme – Language Assessment

Clinicians reported using a variety of static assessments in the hypothetical scenario discussed in the interview. In line with findings from previous survey studies, nearly all clinicians reported that they would use a static norm-referenced test to evaluate the hypothetical bilingual child’s oral language abilities. All but one indicated mentioned using versions of the CELF in some capacity; *“So for the language, I mean, again, we usually use the CELF … I’m trying to think because it’s just so staple. That’s mostly what I would use.”* (P15) and more than half reported they would use versions of the PPVT and/or EVT. *“Um, expressive and receptive vocab, PPVT. Maybe something like an EVT.”* (P13)

When asked about what facilitated their ongoing use of these assessments, clinicians identified several factors, many of which related to environmental context and resources. This included physical resources such as access to testing materials in their workplace; *“Normally it’s the CELF. Because that’s just the one that’s kind of available at our, at our district that we’ve paid for.” (*P27) or the lack of appropriate formal testing materials developed for bilingual populations. *“Standardized testing with any population that’s not the norm referenced population is a little tricky. Again, we have no, no alternative.”* (P22) It also included financial resources. Many clinicians reported that purchasing assessments can be financially prohibitive. *“I would say as well cost is probably a big issue for our team, right? We get only a certain amount of funding that we can dedicate each year to assessment tools.”* (P4) Time was an additional resource factor discussed by several clinicians: *“Uh one because it’s, uh, an easy thing to do. {laughs} It’s really easy to take your CELF off the shelf. {laughs} and it’s harder, and it takes longer to do other types of assessments…It takes more time.”* (P17) and clinicians acknowledged that this is valuable in terms of managing a busy caseload. *“But informal assessments a lot, I think it is, it’s a lot harder to do, takes more time. That’s why we do standardized assessments… because it’s relatively quick and it lends itself to a report and then you can move on to the next kid on your long waiting list.”* (P13)

Beyond environmental and resource issues, many clinicians also identified habit and familiarity as reasons for their ongoing use of these specific NR assessments. *“I mean some of them, to be honest, is just like our board has used them for a long time and like just what we have… I think like honestly out of habit like I think like the CELF is like an assessment that we’ve just had forever…”* (P3) *“Sometimes it’s like really hard to change. So, I think it’s* [the CELF] *something that they’ve always used it, they’re– those are also like I would say–and correct me if I’m wrong, but like the main, or like the typical you know, testing like– it’s the most popular, I would say like test in the whole SLP field.”* (P21) Many clinicians expressed that they continued to use tests like the CELF because it’s what other clinicians in their own workplace do; “*I guess it’s just because it’s the standard that’s being used here, so I guess…I don’t know, this is probably going to sound {laughs}, but if, like it’s what everyone’s doing, so it’s what’s accepted for programming suggestions…I mean, they’re, they’re well used assessments.”* (P9) what clinicians more broadly in the clinical community do; *“I mean, you know, part of it is just they’re widely used in the department, and these are widely used across the profession.”* (P10) and what clinicians have always done. *“We we had a bunch of standard tests that all of us in our department used and basically the CELF, the CELF P, And the various permutations of it as it’s gotten going along, you know, at first it was the CELF, and it was the CELF revised.”* (P12) Clinicians identified that there is value in using a test that everyone knows, because this familiarity means test outcomes are universally understood when reports or other documents are shared. *“Also, they’re selected because when I write about them, when I report about the scores, they’re familiar to other speech language pathologists in the field who might be picking up these reports. It’s very important that they also understand the scores and the the results of the assessment.”* (P14)

##### Subtheme – Literacy Assessment

A different pattern emerged in the assessment of literacy abilities. There was less consistency reported among clinicians regarding what assessments they would use in the hypothetical assessment scenario of a bilingual school-aged child. Clinicians reported using a variety of different screening tools; *“I have a couple of literacy screeners how many I do kind of depends on on the child’s ability, but I I do something called the quick phonics screener which looks at letter and sound knowledge and word reading and sentence reading.”* (P10) including measures like the CUBED which is both dynamic and developed for bilinguals. *“I am learning and loving the CUBED which allows me to look at phonemic awareness as well as that letter-sound knowledge and listening and reading comprehension.”* (P11) Most also report using informal measures either modified from existing screening tools; *“So I I use the quick phonological awareness Screener QPAS, but I usually supplement it with my own stuff… because it doesn’t quite go as in depth as I would like it to… I will also do a um phonics Screener, which I got from this this literacy program I used which is ascend like smarter intervention but I also kind of modified it…”* (P2) or modified from standardized norm-referenced assessments; *“So it started out as Phonological Awareness Test 2, so PAT-2. And then I only used sections of it, and I’ve used it so many hundreds of times that I’ve developed* [it].*”* (P17)

However, most reported modifying the assessments to evaluate skills not addressed in the original measure, rather than for the purpose of rendering it more linguistically and/or culturally appropriate. “*So, for example, there’s a part on the Q Pass for phoneme blending. But sometimes I want to look at more blends like consonant clusters like, can they… blend like a variety of consonant clusters um in initial and final position,” (P2).* Some clinicians report using static norm-referenced assessments, but with much less frequency and consistency than in their assessment of oral language. *“I might do some sub tests of the CTOPP just to get a sense of, of where phonemic awareness is at.”* (P13)

Few clinicians alluded to reasons for this difference in assessment practice patterns between oral language and literacy. Those who did acknowledged that literacy assessment is often a shared responsibility where other professionals, like psychologists, educators or literacy consultants are involved; *“And for the, the literacy part, I don’t do this very often here because it’s the…psychologists that do most of it. And we have also counselors, literacy counselors that do it.”* (P19) Clinicians indicated that these other professionals are often the primary persons tasked with evaluating literacy, while the SLP may play a more of a supporting role. *“We do have a literacy consultant. In a lot of and it’s the line is blurred between like what they do and what we do to be honest, I know that they do a lot more formalized standardized literacy assessments where we do kind of more on the younger side, more, more for yeah, phonemic awareness kind of in that area.”* (P27) Others reported that the scope of practice for SLPs regarding literacy has been changing over time. *“Those of us that have been around a long time, did not do a lot with literacy because we were… we were told you’re not teachers… So, we didn’t do a lot of literacy testing and now I know we’ve got a lot of younger folks in and they’re focusing way more on literacy and they’re using different tools that I’ve not used very much so…”* (P12) Because of this some clinicians report still establishing their literacy assessment practice patterns through professional development or continuing education. *“So, I just recently did in 2021 the Advanced Literacy Practices through UofT, …and that has opened my eyes a lot as to how we could practice differently in our, in our practice as SLPs. Um, I know teachers have taken on a lot, a huge like, historically, they’ve been the only professionals you can support students with literacy needs, but through this program we’ve definitely seen a better role how SLPs can be in the realm of literacy remediation or acquisition of literacy and things like that.”* (P15)

In summary, clinicians report using specific static, norm-referenced English tests (namely the CELF, the PPVT and the EVT) to evaluate oral language skills when assessing bilinguals. Reasons for using these assessments include test availability, financial and time resources, and habit and familiarity. Interestingly, these factors appear to have less influence on literacy assessment. SLPs report using a broad variety of formal and informal tools, including some dynamic assessment, to evaluate literacy and no one tool appeared to be more familiar or habitually used than any other. Reasons for these different practice patterns are occasionally attributed to professional roles and the developing nature of literacy in the scope of SLP practice.

#### Theme 3: Scores reign supreme

##### Subtheme: Language assessment

SLPs reported using a variety of measures in their assessment of bilingual children’s language and literacy skills. This includes those already discussed such as principles of dynamic assessment and modified static NR assessments. Most interviewed clinicians reported consistent use of caregiver and/or educator interviewing with or without an interpreter, *“I would do a really thorough interview with parents and really try to explore that child’s language skills in the home language with an interpreter present.” (P13);* observation of the student using language in context, “*And usually, usually I try to do a kind of an in class observation too, just to get to know them a little bit and see, but that’s usually short maybe 10-15 minutes in the class.”(P27);* and language sampling “*I’m always doing like a speech and language sample for sure, for sure, for sure.”(P16) “I usually start off always with language samples in in both languages.”(P27)* However, as mentioned, nearly all clinicians also tend to use and report outcomes from static NR assessment tools when evaluating oral language abilities. A primary reason identified by nearly all clinicians that perpetuates their use of this practice is the importance of obtaining and reporting a score. *“Well, I need scores, so I often do need assessment scores to help my case.”* (P14)

Scores were reported to be valuable for myriad reasons. Many indicated that they were necessary to ensure children could obtain access to various services including specialized classes in school; *“But then also to get into like a language-based intervention class…they need to show receptive and expressive language scores in a certain range.”* (P9); to facilitate educational identifications or development of individualized education plans; *“If we want to place a child in like a language impairment, they have to be able to score moderately or severe for receptive. And expressive can be mild, moderate, or severe.”* (P4) *“…if down the line identification was, we felt was going to benefit the student often it’s often they’re going off of like more like actual scores versus like that qualitative information…”* (P3); or gain access to intervention or funding. *“So, for kids that we’re doing assessments for the government funding…we need the reports, and our board wants us to write scores.”* (P27) *“If you’re a moderate language delay kiddo, then you get a different pot of money.”* (P7)

There was a perception from most clinicians that scores were required by governing bodies like the ministry of education; *“It’s very hard for that student to access supports within the school system without these measures. So, the Ministry of Education requires standardized measures to show and need to, yeah.”* (P14); or their workplace. *“So, we only do standardized testing at the school board, which goes against everything I believe in as a clinician, but it’s something that we need to do because we have like, specialized classrooms. So, we have to get those scores.”* (P21) It was not always clear whether scores were truly required, and if so, who had mandated their use. “*So then in the fall of last year, I contacted many people from Alberta Ed cause I was determined to get an answer … and no one could really give a clear answer. It’s not very clear why or if they do require standardized score.” (*P27). However, clinicians reported feeling that this was the surest way to get a child access to services. *“Some people have gotten audited, and they didn’t put scores and then they had to go back and add scores. Some people have never put scores, and it’s never been an issue… It’s not very clear why or if they do require standardized score, but that’s just like the safe way to get funding.”* (P27)

Conversely, outcomes of informal measures or dynamic assessments that did not yield scores, while considered useful, were not perceived as sufficient on their own to confirm a diagnosis or identification *“We do have ministry codes that are associated with certain difficulties in schools and so those codes can get funding. So, we, I, I do have to be mindful of those, uh the, the government doesn’t recognize dynamic assessments or informal assessments, unfortunately, there has to be something standardized.”* (P22); or to obtain access to support or services like specialized classrooms. “*But there are public places where it’s like no, like I want to score. You know, we need this score, and this is what these scores mean and it’s like for special classrooms or whatever.” (P16)*

Scores were also seen as valuable because they provided a tidy, quantifiable, and objective metric of performance; *“I I think it’s just the it’s human nature that we do the thing that is most concrete and most historic and gives us the most kind of like tidy answers…and messiness is not something that our profession likes… There’s a lot of good that comes out of doing those assessments I find. But as a profession, we like tidy, and when you can get a number, we we do.”* (P6) that many clinicians felt provided credibility to their test outcomes. *“Because they, they, they* [scores] *give a specific number, right? Like it’s quantifiable this, this, this is this is how they performed. And so sometimes there’s some I guess comfort in that, just to say, oh, this is, this is what they this is how they came out and it’s, it’s nothing that I’m saying or not doing.”* (P18) *“Well, I think probably because like we don’t have–I think a lot of SLPs like the, the, the, the…what I feel is that like they like the scores, they like to know like, it’s not just me who’s, like deciding this, you know?”* (P16).

##### Subtheme: Literacy assessment

The need for scores was not as pronounced in the domain of literacy. While some clinicians reported requiring a standardized score or level producing test; *“So then in terms of the literacy, we go back to the, the literacy part, we’re mainly looking at the reading level. So, we’re going to pick our, our assessment tools for the, the reading level and make sure that we have standardized assessment tools to do that.”* (P22) *“There’s some norms out there for fluency measures for specific ages for falls, you know, fall, winter and spring kind of thing that I’ve used to kind of compare that.”* (P11) many reported that scores were not as critical in the literacy domain as there were for language; *“There’s no real scores, there’s no norm, but I don’t think I really need that to show where the student is in the literacy. Like, you’re not really marking literacy by norm data. It’s really, can they do it, or can’t they?”* (P15) *“I guess you’d have to have standardized scores for receptive language and expressive language and then for literacy, it seems to be a bit more subjective we just have to see how many letter sounds they know. Are they at grade level for reading? So, it seems to be what they take for measures is not, you know, a clear cut, whereas if you are trying to place a student into a specialized program, they have like clear cut, receptive language goal scores.”* (P9)

Some clinicians posited that this was because they did not have the authority in their workplace to recommend a diagnosis or identification of literacy difficulty, *“But other than the LI like language, language impairment title or identification, I don’t really have any–like I can’t bring someone forward for a learning disability or you know, because they’re not learning literacy quick enough like I can’t, I can’t bring that up. That would have to be someone else on the team.”* (P9) Others also expressed that scores were not necessary to confirm reading difficulty, given that it is easier to observe. *“Well, because I guess in terms of like their like early literacy skills, I’m not using any standardized assessments I feel like it it is valid because like all these informal screeners I’m doing, they do often line up with like how they’re doing in the classroom in terms of their reading and writing skills.”* (P3) *“Literacy is more obvious, like they can’t read. Language is more subtle.”* (P2)

In summary, clinicians continue to prioritize using static English NR assessments of oral language with bilinguals because these tests produce scores and clinicians believe school boards and ministries require these scores to confer a diagnosis or to permit access to essential services and support. Clinicians report that outcomes from informal or dynamic assessment approaches cannot be used to for these purposes. The trend is different in literacy assessment, where scores are not as heavily prioritized. It is suggested that this may be because literacy difficulty is more easily observed, and clinicians do not typically assess to diagnose or recommend access to services in the domain of literacy in the same way that they do for language.

### Objective 2: Develop our understanding how SLPs conceptualize and employ validity knowledge

#### Theme 4: Caveats and contextualizing

While many clinicians continue to use and often report scores from static English NR tests, particularly when assessing oral language abilities, this does not necessarily mean that these clinicians perceive this practice as valid. When asked about how they decided that the assessments they used were appropriate, many clinicians who reported using these tests indicated that they perceived this practice as inappropriate and inequitable. They indicated that these static NR tests are not typically developed or normed on bilingual children *“…we know that most standardized assessment features are kind of normed on the white, monolingual middle-class child and so that that doesn’t reflect the vast majority of the kids that I see.”* (P23); that the test items may not be linguistically or culturally appropriate “*They’re not appropriate for the kids in the North We have no escalators. There are no alligators, there are no–you know the vocabulary and what’s relevant to, in assessment is going to be different for a southern child.”* (P17); that these tests may disadvantage children with different linguistic backgrounds *“I think it disadvantages children of other languages, other language, and cultural backgrounds. They’re disadvantaged when we give them our standardized tests.”* (P12); and that they do not necessarily accurately represent their abilities *“Well, these tests usually are normed on monolingual speakers of the language or the test that the language is, is checking for so I don’t think that represents the profile of the kids very well.”* (P25)

To contend with the perception that scores from static English NR tests are required to grant children access to services and their own beliefs that these same tests may not be suitable for use with bilingual students, most SLPs reported using caveats or disclaimers when reporting static English NR scores in bilingual assessment. *“So I would, I would proceed in English but with sort of the uh caveat in the report that English isn’t the first, the student’s first language and we have a standard proviso in the report regarding that that you know the score should be taken well, not with a grain of salt, that’s not the language, but should you know, be considered in light of the fact that the student’s first language and that the norms are based on native English speakers.”* (P10) *“I also a lot of the times I my conclusions are always have like a little disclaimer somewhere in …because we, you know, such and such is bilingual or trilingual, this assessment was made in this language. So just keep in mind that it doesn’t reflect the reality of, you know, his, his level in in general, so there’s always disclaimers everywhere in my reports.”* (P19)

Several clinicians acknowledged the limitations of using caveats, saying that even when one was included, it did not always deter readers from focusing on the scores. *“I mean, obviously it’s norm referenced against English speaking children, monolingual children, so it’s obviously not, not a true true norm reference because it’s not the population that we’re using it with. But we do put in our report if they are bilingual like, test scores should be used with caution. They are norm referenced against monolingual children, so there’s like a disclaimer twice in our reports that we have to write if they are bilingual, um, just to that, but I feel like it’s also, you know, you read that, but then you also immediately look at the test scores so…”* (P9).

A small number of interviewed clinicians had started to abandon using caveats and are considering how they can change their assessment practices. This includes using adaptive scoring procedures with bi-dialectal and bilingual students *“And I would say for the first 5 1/2 years or so I went along with the little disclaimer on the top of the assessment and saying, you know, this kid comes from non-English speaking background and read this with caution and hesitancy and take with a grain of salt, blah blah blah… So, I get them to say it in their language and I write it on the side. So, I’m like that doesn’t necessarily mean that doesn’t count. They know what the word for umbrella is. They just couldn’t tell me in English. They haven’t been here long enough, or they weren’t exposed long enough to English in at home to say the word umbrella. Am I going to score them as a zero? No. I think that’s wrong.”* (P15); leveraging their own cultural and linguistic knowledge to adapt existing tools *“I have had translated the Child Development Inventory, so MacArthur Bates, I’ve had that translated into standardized Inuktitut.”* (P17); and relying more on less linguistically biased alternatives like nonword repetition tasks *“It’s repetition of nonsense syllables. So, you say like Tay toe and the kids say Tay toe and then you say Tay Ta Ta and you see how many syllables they can do. And the kids that truly have language disorders. They really can’t do it. They can’t hang on to that.”* (P12); or dynamic assessments to inform their decision making and to include in their reporting *“Sometimes I also do…for younger kids, so the cube has the dynamic decoding measure… I always do a nonword repetition test just to add in to my screen… I’ve like to read and know through research that is like helpful to just differentiate.”* (P7).

In summary, clinicians who use static English NR tests developed for monolinguals with bilingual children do so for various reasons. Their use of these tools does not mean that clinicians perceive this practice as valid or appropriate. Clinicians know that these measures may not accurately represent a bilingual child’s true ability. They communicate this by reporting caveats and disclaimers alongside test outcomes. Some are starting to move away from reporting scores and caveats and are exploring other adaptations and alternatives.

#### Theme 5: Clinical knowledge vs. System constraints

##### Clinical knowledge

Making valid assessment choices extends beyond choosing a test designed to be used with the population assessed based on their linguistic characteristics. Other factors, such choosing a test that is designed to be used for the purpose of the assessment; that has adequate psychometric properties; whose outcomes can provide insight into real life performance and whose results can be used to make decisions or act with minimal risk of adverse outcomes are all critical to consider (Kane, 2006).

Several clinicians alluded to psychometric properties when asked how they decided whether tests were appropriate to use. Clinicians usually only alluded to “reliability and validity” as opposed to specific properties like sensitivity and specificity, standard error of measurement or parallel form reliability when discussing how they decide whether tests were appropriate for use. *“We read through the manual and there would be the extensive research that was done, the validity, the reliability and it’s pretty stock standard.”* (P15). Many reported not feeling confident with their knowledge of the different properties and their implications for valid assessment practices. *“To be really, really honest I don’t even I, I don’t have the, the knowledge to really critically look at the reliability and validity data…”* (P13). Other clinicians indicated that psychometric properties were not a top priority when selecting a test. *“…like I know the TILLS and the CELF have psychometric properties that-again this is something that you probably are going to hear this a 1000 times. Like, I’m not like studying the psychometric properties. I’m more so going off the trust of the SLP community and determining that these are like well-used tests for a reason.”* (P2) Finally, some clinicians also considered tests to be valid if they had been developed by researchers or academics *“…I trust the researchers who said these were, you know, valid measures and everybody else in my department is using them.”* (P10); regardless of their properties *“Um, I skim that section on reliability and validity… I don’t think I’ve ever seen a manual like, for example, a CTOPP manual or a CELF manual, one of those big, big tests where they…report having any concerns… I think I do think that we take it on faith with those big tests. We take it on faith that they’ve, because they’re academics, we’ve developed them.”* (P13).

Some of the clinicians interviewed felt reliability and validity were important factors to consider when deciding if a test was appropriate, but several reported limited knowledge of these psychometric properties. Others felt that these properties were not the most important factor informing assessment selection. Clinicians reported trusting that researcher-developed tools commonly used by their clinical peers were valid and appropriate.

##### System Constraints

When discussing their approach to the hypothetical assessment scenario, few clinicians identified what their purpose for assessment would be without prompting. Identifying the purpose of an assessment is a key factor in making valid choices about which assessment measures to use. When prompted about why they assessed, their purpose or reason for assessment appeared to be constrained to their practice context. In public contexts, like schoolboards, the primary purposes for assessment were typically identifying difficulty/diagnosing *“Well, I’m trying to gather as much information about the student’s strengths and weaknesses in terms of communication, to the best of my ability, and then once I figure out the areas of I don’t want to say areas of weakness, but areas that are in need of of strengthening then I can give a good picture about, you know, whether or not this looks delayed or potentially disordered and the severity.”* (P14); determining eligibility for Individual Education Plans (IEPs), *“The school board is kind of both. They want, you know, less goals, unfortunately, more like what did the outcome say because they will go off and do an IEP.”* (P15); accessing funding “*then I would do a formal assessment, because usually, at least in Alberta, for funding, we need that formal or standardized assessment to get services and funding in the schools.”* (P25); or obtaining access to specialized classes “*So, but we’re the ones, you know, who are first figuring out does this seem like an LI kid to me? Like an LI class kid to me.” (P10).* Many schoolboard SLPs reported that their workplace restricted the parameters of practice *“…when I’m in the school board, you know, because it’s a public setting sometimes you’re kind of stuck with like what can I and can I not do, you know? Because it’s not just a question of what can I do as an SLP, but what can I do as a school board employee.”* (P16).

Public clinicians also reported that providing direct therapy was often not an option in their workplace. *“And if you know if it were a model where we could do intervention, then what I would be doing is looking for the goals for me to work on with that student. But it’s that’s not the way it works…” (P10).* Assessing to determine treatment strategies or recommendations was also not always seen as the most effective use of time, given that many reported that there was minimal capacity in the system for follow through on these recommendations *“In education, often, yeah, what we found is you go through the lengthy process of assessing and reporting and then the report goes into a confidential file and it’s not read by, you know, next year’s teacher or, yeah, any of the professionals who would be able to provide support because everybody’s spread really, really thin.”* (P11); meaning that even if they assessed for and included this type of information, they did not feel it would be likely for it to be used *“It’s sort of the existential the underlying, unspoken, existential question there is who’s going to use that information?… I actually think that is probably the real, the real issue, is that we all know nobody is going to use that information.”* (P13).

Conversely, clinicians in private contexts were more likely to report assessing to set treatment goals *“Whereas private practice would be more like direct like you’re doing it for your, you know, for the parents, but also for yourself to try and see where you could go with therapy.”* (P9); or plan intervention *“I don’t know if it’s different in the public world, but in the private world like when, when families are coming to like request an assessment, there typically is the intention to continue with therapy if it is warranted We see that there’s some difficulties, like we consent to continue starting with therapy and we’ll kind of develop that therapy plan.”* (P23). Private clinicians also reported feeling that they had much more flexibility in the types of assessments they could use. *“So, another really great thing about being in private practice is that I have a lot of flexibility with how I do my assessment And so, I really do, even though I try to use standardized assessment, I really, I feel like I supplement a lot more with informal tools and I’m able to do so because I have a lot more flexibility in my practice…relative to the public system” (*P23).

However, there were still system level issues that affected private practice assessment patterns. Often clinicians were more likely to prioritize assessing to set treatment goals without first diagnosing or confirming difficulty, because coming to private therapy alone was perceived as indicative of a problem, *“Nobody comes and seeks out private practice unless they know there’s a problem. So, there’s already been a problem identified whether it’s diagnosed or not. I’m literally just assessing for the purpose of informing my intervention and recommendations.”* (P2). Jumping right to setting treatment goals without a diagnosis or comprehensive assessment of abilities, was a way for clinicians in private practice to save time*, “Um* [redacted private clinic] *is a place for therapy, so the assessment is an assessment to treat. It rarely goes from interview to testing. It usually goes from interview to therapy because the families at* [private clinic] *are by definition seeking out therapy for a particular reason.”* (P6); and to save their clients’ money *“…private SLP is expensive. A lot of people don’t have benefits. A lot of people are paying out of pocket.”* (P23).

The difference in purpose of assessment based on practice context is summarized neatly by clinicians who had experience working in a public and private practice. *“Because in public they want an identification and private they just want help.” (P2)* This difference directly affected how clinicians operated in these two contexts. *“Who I work with will decide what the purpose of this assessment is, so if I’m working for* [redacted agency name] *they are nonprofit, they’re and it’s FASD assessments. So basically, I am doing the assessment so that I can contribute to the FASD team. Whereas in the Western Arctic I work with the kids, and so for those assessments, it’s really for choosing therapy goals.”* (P17).

The purpose of assessment appears to be defined by practice context to the extent that clinicians often did not articulate the objectives of their assessment without prompting. It is implied that most public SLPs assess to identify and determine if there is difficulty, while most private SLPs assess to inform intervention.

In summary, clinical knowledge of valid assessment practices must be considered both through individual clinical level knowledge, but also through system level constraints that affect a clinician’s ability to make valid assessment decisions. At the individual level, clinicians report limited knowledge of psychometric properties and do not consistently use this information to decide whether a test is appropriate for use. However, even when clinicians are aware that an assessment practice is inappropriate, the systems they work in often constrain their assessment practices. This is evidenced by the many clinicians who reported knowing that using a static English NR test to evaluate a bilingual child’s language abilities is inappropriate, but who continued to do so anyways, because the systems in which they operated required that they produce diagnoses and test scores.

## Discussion

This qualitative study examined the clinical decision-making processes that informed assessment practices of clinical Speech-Language Pathologists (SLPs) in the evaluation of bilingual children’s language and literacy skills. It explored what factors affected use of different types (e.g., static vs. dynamic) of assessments, and how clinicians conceptualize and employ validity knowledge in their choice and use of assessments. We begin by addressing the limitations of this work.

### Limitations

First, despite our best efforts, we were not able to recruit clinicians from the Eastern provinces to participate in this study. We were also only able to recruit one participant from Québec. Given that education and healthcare are provincially and territorially mandated in Canada, systems and processes that govern assessment practices may differ. Furthermore, patterns and profiles of bilingualism also vary across provinces and territories. For instance, the most common bilingual profile in Québec and the Maritimes is English and French, but the most common bilingual profile in British Columbia and Toronto, Ontario is English and Mandarin (Statistics Canada, 2022b). Consequently, it is possible that clinical perceptions of factors affecting assessment practices with bilinguals may differ across Québec and the Eastern provinces, and that these perspectives are not represented here. Secondly, all but four of the twenty-one clinicians we interviewed were bilinguals (80%). Nationally, only 40% of the population is bilingual (Statistics Canada, 2022b) and within the SLP population, rates of bilingualism are lower, though no nationwide data has been collected to establish this. It is possible that the interviewed clinicians self-selected to participate in a study about bilingual assessment practices because of their own linguistic profiles. Monolingual English-speaking clinicians may have different perspectives on bilingual language and literacy assessment practices.

### Clinical conceptualization of validity in assessment

When clinicians were asked what type of assessments they used, they mentioned a variety of different types of assessment measures including but not limited to static English NR measures, dynamic assessments and modified or adapted versions of static assessments characterized here as the process of “assessing dynamically.” We consider the clinical decision-making processes that inform the use of each of these types of measures, and the clinical understanding of valid use of these assessment types.

#### Static Norm-referenced Assessment

Consistent with findings from previous survey studies, we find that many clinicians continue to use static norm-referenced assessments developed for English monolinguals, with bilingual school-aged children (Arias & Friberg, 2017; Caesar & Kohler, 2007). Clinicians often mentioned ease and time required for administration, familiarity and clinical reputation and cost and access to tests as factors influencing their choice of assessment. When discussing how they decided whether these assessments were appropriate or valid for use, few clinicians mentioned psychometric properties as a factor in selecting an assessment. Most reported not feeling confident in their knowledge of test psychometrics and did not mention any specific property as influencing their decision. This is consistent with findings from prior survey studies (Kerr et al., 2003). Many clinicians reported that if assessments were researcher-developed and well-established in the clinical community, that they were likely valid for use. Very few clinicians mentioned how the purpose of their assessment would inform whether a test was appropriate to use, despite this being a critical factor in making valid assessment choices (Kane, 2006). In line with prior studies, results suggest individual clinical level knowledge of validity in assessment can continue to be improved.

The need to increase individual clinical knowledge of psychometrics has already been discussed in previous literature, and in our opinion is not the most salient finding from our interviews or the most pressing factor to consider in terms of improving valid assessment practices with bilinguals. One factor that clinicians identified consistently as heavily influencing their test selection is whether a measure produced a score. Through our in-depth conversations with clinicians, we learned that the score itself may not be inherently valuable to the clinician. Indeed, many clinicians mentioned that they felt the score might not necessarily reflect a child’s true ability and that results from static NR tests did not truly represent bilingual children’s skills. Rather, the power of the score lay in the outcomes that it allowed a child to access. These included key educational and therapeutic services like diagnoses, individual education plans and access to intervention.

From Messick’s perspective, these clinicians could be conceived of as prioritizing the *consequential validity* of their assessment choices (1989). Consequential validity relates to the potential positive or negative value implications (e.g., accurate or inaccurate diagnosis) and the potential actual consequences (e.g., access or no access to services) that can arise from assessment outcomes (Messick, 1989). The stakes of clinical language and literacy assessments for children are high. It is possible that clinicians are weighing the risk of misrepresenting a bilingual child’s ability using a static norm-referenced test (a negative value implication) against the risk of not being able to provide a child who might need it access to key services with a score from a static NR measure (a negative actual consequence). If a score is required or perceived to be required, and there are no score-producing tests available for that child’s language profile, the clinician is between a rock and a hard place in terms of making the decision that results in the least amount of risk for their client, and the most positive potential actual outcomes. While we do not advocate that using English static NR assessments with bilinguals is the appropriate response, we contend that teaching individual clinicians about psychometric properties and other aspects of validity in assessment will not resolve this issue. Rather, systemic changes to how services are granted to children along with development and increased access to more appropriate assessment materials for bilingual and CLD populations will be required.

#### Dynamic assessment & Assessing Dynamically

Consistent with findings from surveys studies conducted by Caesar and Kohler (2007), Arias and Friberg (2017) and D’Souza et al., (2012) that investigated frequency of use of dynamic assessment with bilinguals we find that few of the interviewed clinicians report using “dynamic assessment.” However, most interviewed clinicians did report using principles or approaches that could be considered dynamic in nature like prompting, teaching, providing feedback and repetition and re-evaluating performance in testing. We contend that there are several reasons why clinicians might use “dynamic assessment,” in some instances, and “assess dynamically” in others.

First, this practice pattern could be due to lack of clarity around the definition of “DA.” Historically, a main criticism of DA has been its conceptual fuzziness (Caffrey, 2008). There are many different types of DA, which vary based how closely they approximate treatment (e.g., as in interactionist DA that seeks primarily to remediate difficulty) vs. evaluation (e.g., as in interventionist DA that seeks primarily to assess; (Lantolf & Poehner, 2008). Clinicians who “assessed dynamically” recognized that this practice was not consistent with evaluation the way static, norm-referenced assessment might be. However, they also did not describe it as “dynamic assessment,” suggesting a lack of certainty around what DA is and isn’t. Providing a comprehensive definition of DA is beyond the scope of this paper, though we believe it is an important avenue for further exploration. Importantly, even if these clinical practices are not described as textbook “dynamic assessment” by SLPs, our findings suggest that many clinicians do have the skills and knowledge to incorporate “principles of dynamic assessment” in their evaluations.

A second possible reason why clinicians report using both “dynamic assessment” and “assessing dynamically” may be because of a lack of tools, evidence and guidelines on the use and application of DA in the varied complex clinical situations where it may be required. For starters, clinicians who reported using “dynamic assessment” typically referred to a specific, commercially available dynamic assessment, such as the CUBED dynamic decoding measure, while those who reported “assessing dynamically” often mentioned making adaptations to existing commonly used assessments (e.g., incorporating prompting, teaching and re-evaluating in the CELF-5). This latter practice could be characterized as adaptive expertise (Cupido et al., 2023). When engaging in adaptive expertise, clinicians draw on their conceptual knowledge to build new ideas, innovate to solve problems, and create work-around or new strategies to contend with practice issues that cannot be solved using existing or available approaches (Cupido et al., 2023). In most practice situations a clinician encounters, there is not a “dynamic assessment” available for clinicians to use, or concrete guidelines or how to integrate a DA approach. Through the lens of adaptive expertise, clinicians who are confronted with these complex practice situations use what they already know and have access to (e.g., static NR English tests) and adapt to meet the needs of their practice (e.g., incorporate principles of DA by adapting or modifying static tests for bilinguals). Conversely, clinicians who use “dynamic assessments” would do so because there is a readily available assessment measure that meets the requirements of their practice and does not require adaptive expertise to implement. As an example, the CUBED is commercially available, free and developed by researchers for use with bilingual students. It is also relatively quick to administer and produces normed scores that, as discussed above, are perceived as necessary for accessing certain outcomes for clients.

#### Implications

##### Clinical education in dynamic assessment

Given that there appears to be uncertainty around what is and is not dynamic assessment, and the purposes it can serve in practice, it would be valuable for clinical education programs to provide increased instruction regarding theoretical underpinnings of various dynamic assessment approaches and the potential of DA to not only inform treatment goals, but also to help differentiate between difference and disorder in bilingual populations. Beyond teaching about DA, we also contend that teaching clinical students the value of adaptive expertise will be essential. Three key practices to developing this expertise identified by Mylopolous et al. (2018) in clinical health professional include (i) emphasizing understanding in learning (over recall), (ii) providing students with opportunities to encounter complex problems and apply their learning to make discoveries, and (iii) maximizing the variation in instruction of clinical concepts. Our findings lead us to argue that it would be most valuable to frame these complex problems in clinical education through the lens of problem-posing education (PPE) as proposed by Freire (Houser, 2007). In a PPE approach students are presented with clinical cases, and encouraged to consider not only the biomedical individual level factors (e.g., clinical profile indicative of reading disorder as a result of a known medical condition like dyslexia), but also to interrogate the social and political contexts in which their clients are situated (e.g., the inequities of the education system responsible for allocating reading assessment and intervention services). In this approach, clinical students are trained to recognize individual and systemic origins and influences on disorders and are poised to advocate for changes (Cavanagh et al., 2019).

##### Dynamic assessment tool development

For researchers engaged in development of dynamic assessments, results from these interviews and previous survey studies suggest that habit, clinical reputation, and familiarity are factors that clinicians prioritize when choosing a test. To capitalize on this, it may be useful to examine whether modelling new dynamic assessments after existing static tests with established reputations or developing modified dynamic versions of these tests will result in increased clinician uptake. Modelling DAs after existing SAs may also minimize barriers to uptake related to time required to learn new assessments and integrate them into practice, another factor identified by clinicians as relevant in selecting which assessment to use. While percentiles and standard scores that compare a child’s performance to their peers is not consistent with a dynamic assessment approach, uptake of DAs may be more likely if they produce some type of score or numerical representation of ability, particularly for tests evaluating language, as this was a factor many clinicians identified that perpetuated their use of SAs, particularly in school settings. In new DAs, this could be as simple as a raw score and a corresponding label (e.g., developing vs. developed skills) rather than a scaled or standard score. In the long-term we advocate for clinical education to prepare clinicians to question systemic practices such as these. However, whether equitable or not, scores are still required or perceived to be required in many education and healthcare settings. These scores are used to categorize, characterize ability, and easily transfer information across professionals and settings. In the short-term, a score-producing DA could be valuable in these contexts.

##### Addressing organizational barriers to dynamic assessment

The outcomes of these interviews indicate that many clinicians continue to engage in assessment practices they perceive as invalid or inappropriate (such as using static NR English measures with bilingual children) because the systems in which they practice require this information to provide these children access to essential services. Our findings that SLPs’ assessment practices are influenced by more than their individual clinical knowledge of validity is critically important. Future research efforts that seek to shift assessment practices need to consider the reasons why SLPs engage in assessment practices and identify the appropriate targets for practice change in specific contexts (Damschroder et al., 2022). For instance, if there are codified policies in schoolboards or ministries of education requiring SLPs to report norm-referenced scores for a child to access services, then advocacy efforts may be appropriate to address this system level barrier. In this context, SLPs can advocate on behalf of their clients for use of alternative less biased, linguistically, and culturally appropriate measures like dynamic assessment, modified or adapted versions of static assessments, language sampling and informal observations. Clinicians can also question and interrogate why potentially biased scores from norm-referenced measures are required for children to access critical educational services. Regulatory bodies in Canada, like SAC could support clinicians in these efforts by advocating for clarity from provincial and territorial ministries of education regarding whether and why scores are required in certain practice contexts. From a research perspective, examination of whether scores are necessary in school contexts, and why these numerical representations of children’s ability are so ubiquitous in healthcare and educational settings is also a promising avenue for exploration. More broadly, further investigation into specific factors affecting assessment practices in different clinical contexts (e.g., publicly funded preschool programs), across different domains of practice (speech, voice, fluency etc.), and with different populations (e.g., based on age or disorder profile) will inform what actions clinicians, educators and regulatory bodies can take to continue to promote valid and appropriate assessment practices across the spectrum of clinical practice.

## Conclusion

Clinicians weigh multiple factors when choosing which measures, dynamic or otherwise, to use when assessing bilingual school-aged children’s language and literacy skills. Like prior work, we find that these decisions are influenced by practical factors like test availability, time and resources and by clinical knowledge of test psychometrics and valid assessment practices. However, we also find that these choices are influenced by dominant biomedical discourses and workplace practices and routines. We contend that practices that may have previously been considered test misuse, such as adapting static assessments, be reframed as adaptive expertise that draws on appropriate validity concepts such as consequential validity. When encountering a situation like the assessment of a bilingual child for which there is limited available evidence or resources, clinicians creatively adapt and generate novel solutions to meet the needs of their clients and produce the best possible outcomes.

## Supporting information

Supplemental – Interview Guide

## Acknowledgements

First, we want to acknowledge the clinicians who participated in this study. We are grateful to them for sharing their time, perspectives, and opinions. This study was supported by an Ontario Graduate Scholarship from the Ministry of Colleges and Universities, a Doctoral Canada-Graduate Scholarship from the Social Sciences and Humanities Research Council of Canada, and a Duolingo dissertation grant in literacy awarded to E. Wood; as well as a Social Sciences and Humanities Research Council of Canada Insight Development Grant awarded to M. Molnar.

## Author contributions

**EW** Conceptualization, methodology, data collection, analysis, writing, revising, funding **MK** conceptualization, analysis, writing, **OD** conceptualization, methodology, revising, **MM** conceptualization, methodology, revising, supervision, funding

## Data availability statement

The interview guide can be found in the appendix.

