## Supplemental – Interview Guide for "Why did you use that test? Exploring speech-language pathologists clinical decision-making in bilingual language and literacy assessment"

**Semi-Structured Interview Guide**

**CONSENT REVIEW**

*Thanks for agreeing to talk with me today. I know how busy you are, so I really appreciate you making the time.*  *Before we begin, do I have your consent to audio and video record this conversation? The recordings will be housed in a password protected folder on a secure U of T platform.  Only members of the research team will have access to them. We will save the recordings with a participant code, so that the file cannot be linked to your name. The recordings will be deleted when they have been transcribed and analyzed. Do you have any questions about the recordings?*

*Ok, thank you so much. I also want to remind you before we begin that you are free to withdraw your consent to participate at any time. We can also stop the interview at any time at your request. If you are interested in learning about study outcomes, you are free to email me following the interview, and any products of the research, such as a research article, would be shared with you then. Do you have any questions for me about this?*

**BACKGROUND REVIEW**

*Great, so first, I would like to tell you a bit about me, and then get a better sense of your work experience. So, I am a clinical speech-language pathologist. I have been working for about 8 years now. For this study, I am interested in learning more about how SLPs make decisions in the assessment process. There is a lot of research out there about what we do, but not all of it gets at why, or explicitly asks us as clinicians. Any questions about that?*

*Alright, and I know you reported some of this information on the background questionnaire, but can you give me a brief background of how long you have been practicing, what population you have worked with and where you have practiced?*

**QUESTION 1 – ASSESSMENT OBJECTIVES, TYPE AND QUALITY OF TOOLS USED**

*Thank you. Now I am hoping we can talk about a hypothetical situation. Imagine you are at your current workplace, and you have received a new referral for an assessment of language and literacy skills.  The child is a seven-year-old, male, bilingual student in the second grade.  Can you walk me through your assessment process as a clinician? What do you do?*

**Probe - Objectives**

**If objective not mentioned - probe for objectives**

- *In your experience, what might the objectives of such an assessment be?*

**If/after objective mentioned/discussed - probe for assessment measures**

- *So, if your objective is ____ what assessment measures would you usually use in this type of scenario?*
- *How have you typically decided that those measures are high quality?*

**Probe - Dynamic Assessment**

**If only static assessment measures mentioned probe for dynamic assessment**

- *You mentioned using ____assessments. Are there other types of assessment measures you would use?*
- *Have you used dynamic assessments?*
- *If not, what has prevented you from using dynamic assessments in clinical practice with bilinguals?*

**If dynamic assessments mentioned probe for experiences and facilitators**

- *Can you describe your experiences with dynamic assessments?*
- *Do you regularly use them? If yes, what has facilitated this?*

**QUESTION 2 – INTERPRETATION OF FINDINGS AND OUTCOMES**

*We have discussed some of the different types of measures one might use in this assessment scenario, and the ways in which you would determine whether those measures were appropriate or high quality.*

- *Once you have completed the assessment and are interpreting your test outcomes, how do you integrate findings to draw conclusions?*
- *What happens after you complete and share your assessment findings?*
- *What decisions are made based on and what are the implications of these outcomes?*

**QUESTION 3 – USING NR ASSESSMENTS WITH BILINGUALS**

*We know it is common for SLPs to use and interpret/report norm-referenced standardized assessment measures with bilingual students.*

- *How do you see this issue?*
- *Why do you think this practice persists?*

**QUESTION 4 – IDEAL ASSESSMENT SCENARIO**

*Now we have discussed some of the things that make it easy to carry out certain assessments and challenging to carry out others.*

- *In an ideal world, what assessment measures would be available to you that would ensure you could provide a high-quality assessment for this child?*
- *In your opinion, why don’t these types of measures exist?*

**WRAP-UP**

*That brings us to the end of my questions. Thank you so much for your time and insights today. Do you have any questions for me?*
